# North Carolina CORonavirus VAriant SEQuencing (CORVASEQ): A surveillance network to monitor an evolving pandemic

**DOI:** 10.1101/2025.11.25.25340988

**Authors:** Bethany L. DiPrete, Audrey E. Pettifor, Justin Lessler, Corbin D. Jones, Jennifer Anderson, Amir Barzin, Amy James Loftis, John T. Fallon, Melissa B. Miller, Jeremy R. Wang, Thomas N. Denny, Miranda Carper, Cynthia Gibas, Jessica Schlueter, Gregory A. Hawkins, Kimberly D. Reeves, Christopher M. Polk, Mindy Marie Sampson, Angela Harris, Erica F. Wilson, Zack Moore, Jeffrey Warren, Blossom Damania, Dirk P. Dittmer

**Author notes:** Corresponding Authors: (Dirk P. Dittmer) and (Blossom Damania).

## Abstract

When the need for comprehensive viral genomic sequencing emerged early in the SARS-CoV-2 pandemic, public health leaders in North Carolina sought to expand the state’s genomic surveillance capacity. The North Carolina CORonavirus VAriant SEQuencing (CORVASEQ) network was established in the summer of 2021, drawing on institutional and local partnerships to build a statewide pathogen genomic surveillance network to conduct timely and representative genomic surveillance of SARS-CoV-2 variants. This streamlined and comprehensive network rapidly increased the number of viruses sequenced and provided representative genomic surveillance coverage of all 100 counties in North Carolina. The standardized protocols and established modes of communication developed in establishing the CORVASEQ network can help the state more rapidly mobilize in response to ongoing and emerging public health threats in North Carolina. The successes and lessons learned from the CORVASEQ network can serve as a valuable roadmap for other states or areas as they seek to build or enhance pathogen genomic surveillance networks.

## Introduction

When SARS-CoV-2 was declared a global pandemic in early 2020, public health systems faced unprecedented surveillance and pandemic response challenges. After the first whole genome sequences of the novel SARS-CoV-2 virus were published in January 2020 (1,2), surveillance efforts initially concentrated on developing and disseminating diagnostic RT-PCR and rapid antigen tests. Increased access to rapid antigen and PCR testing allowed for the monitoring of case counts, hospitalizations, and deaths attributable to COVID-19. As the virus evolved and new variants of concern (VOCs) began to emerge, genomic surveillance quickly became a critical tool in tracking the evolution and spread of SARS-CoV-2 (3,4). Timely genomic surveillance provides a deeper understanding of viral dynamics in near-real time than can be ascertained from case monitoring alone, allowing researchers and public health professionals to track the emergence and spread of pathogen variants.

Coordinated, rapid sequencing efforts proved essential in understanding SARS-CoV-2 transmission dynamics and disease burden throughout the pandemic. Several countries quickly mobilized national networks to ramp up sequencing efforts in response to emerging variants of SARS-CoV-2. Among the most notable was the United Kingdom, which developed a nationwide consortium for genomic sequencing of SARS-CoV-2, in addition to research and training efforts, allowing the country to achieve impressively high sequencing volumes (5). Significant capacity-building efforts for pathogen genomic surveillance were also conducted in several African countries during the height of the pandemic. In particular, the South African Network for Genomics Surveillance of COVID (SANGS_COVID) was formed in May 2020, which allowed South Africa to scale up its sequencing capacity. These efforts proved instrumental later in the pandemic when Omicron (B.1.1.529) was discovered in SARS-CoV-2 genomes from South Africa and Botswana (4,6,7). This early warning signal allowed the rest of the world to prepare for the global spread of this new, highly transmissible variant. In the US, central sequencing efforts were augmented by academic medical centers, which had the capacity and domain expertise to handle infectious viruses and conduct next-generation sequencing (8–10).

Collaborative networks have proven fruitful for the surveillance and study of other pathogens across geographies and populations. PANGEA is a research network focused on using viral genomics and epidemiologic data to understand HIV transmission in sub-Saharan Africa with the goal of informing targeted prevention efforts (11,12). The HIV Prevention Trials Network (HPTN), HIV Vaccine Trials Network (HVTN), and AIDS Clinical Trials Group (ACTG) were established to expand the evidence base around global HIV prevention and treatment. Since their conception, studies from these networks have significantly contributed to our understanding of HIV transmission, effective treatments, and prophylaxes for combating the HIV pandemic (13–20). These networks were successfully able to mobilize to test vaccines and other therapeutics for COVID-19 as part of the Coronavirus Prevention Network (CoVPN) (21). The CDC has established networks for monitoring seasonal influenza and vaccine effectiveness, such as the US Flu Vaccine Effectiveness (VE) Network, VISION VE Network, Respiratory Virus Transmission Network for Influenza (RVTN-Flu), and the Investigating Respiratory Viruses in the Acutely Ill (IVY) Network, with the latter having also expanded to study COVID-19 vaccine effectiveness. Finally, there are national surveillance systems managed by CDC and state health departments, including ArboNET, which monitors human arboviral infections, veterinary disease, and vector populations (22), and PulseNet, which monitors outbreaks of foodborne illness (23).

While establishing coordinated sequencing networks can strengthen surveillance and analytic capacity, developing surveillance protocols and networks is time and resource intensive. This challenging task is further complicated during outbreaks when public health resources must be concentrated on responding to a given public health emergency. It is thus crucial to establish coordinated and comprehensive surveillance networks that can be rapidly mobilized in times of crisis. To improve statewide surveillance capabilities, the North Carolina Department of Health and Human Services (NC DHHS) partnered with the North Carolina Collaboratory to form the CORonavirus VAriant SEQuencing (CORVASEQ) Surveillance Program, drawing on institutional and local partnerships (24) and expertise to build a statewide pathogen genomic surveillance network. The primary goal of this network was to form a comprehensive surveillance system to conduct genomic sequencing representative of SARS-CoV-2 infections in individuals across the state. Its secondary goal was establishing a network that could be mobilized in future public health emergencies using standardized genomic surveillance protocols.

## Methods

### Establishing the North Carolina CORonavirus VAriant SEQuencing (CORVASEQ) Surveillance Network

The North Carolina Collaboratory is a dynamic research center established in 2016 by the North Carolina General Assembly to facilitate and promote research activities across the University of North Carolina (UNC) system and other academic institutions across the state. Research activities span a wide range of disciplines, the findings of which are then communicated to state and local government officials to translate into policy and action. Thus, it was the logical choice to lead and administer CORVASEQ.

The Collaboratory has funded over 90 other projects related explicitly to COVID-19. When the need for comprehensive sequencing emerged, the Collaboratory and NC DHHS sought to expand genomic surveillance capacity in North Carolina. Prior to the fall of 2021, pathogen genome sequencing in North Carolina was mostly limited to sequencing facilities at major academic institutions and hospitals, limiting the ability of public health professionals and researchers to conduct representative pathogen surveillance throughout the state. CORVASEQ was established in the summer of 2021 with the goal of creating a streamlined network of institutions and laboratories to conduct representative genomic surveillance of SARS-CoV-2 variants and communicate findings to NC DHHS and state public health leaders in a systematic and timely fashion.

The CORVASEQ network is comprised of *sample acquisition partners*, *sequencing partners*, and the *epidemiology data core team* (**Figure 1**). Specifically, the sequencing partners include Duke University, East Carolina University, UNC-Chapel Hill, NC State University, UNC-Charlotte, and Wake Forest University. Sample acquisition partners who provided COVID-19-positive samples for sequencing from all 100 counties in the state included the healthcare systems affiliated with Duke, UNC, and Wake Forest, as well as the following healthcare systems: Atrium Health, HCA Healthcare, Novant Health, WakeMed Health & Hospitals, the Veteran’s Affairs (VA) System, and ECU Health. The sample acquisition partners represented not only tertiary hospitals but also community hospitals, health clinics, and physician offices.

**Figure 1.**
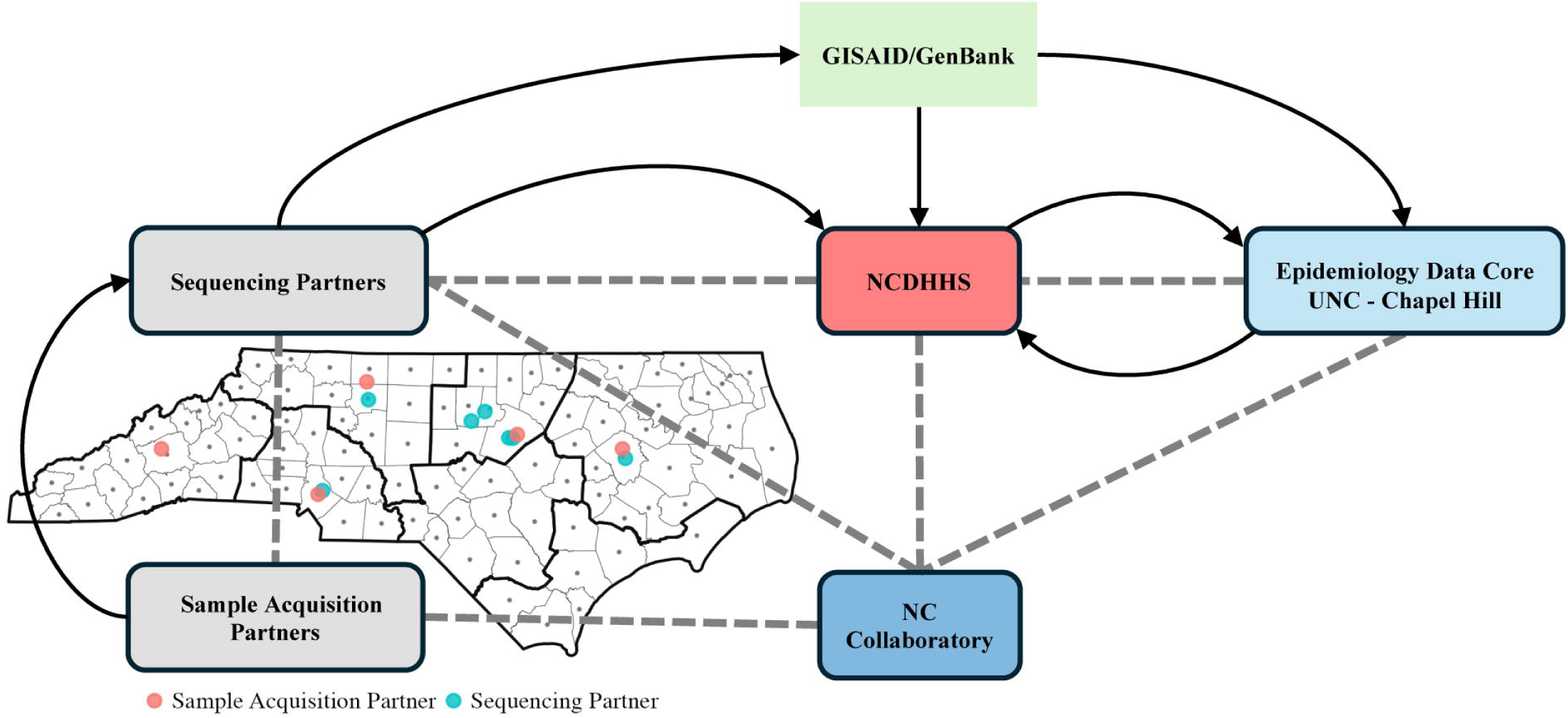
Network structure and flow. Black arrows indicate data transfer, dashed lines indicate regular communication. Sequencing was completed in all 100 counties, as indicated by black dots. Geographic outlines indicate flu regions for monitoring seasonal respiratory pathogens (as defined by NCDHHS).

### Sampling, Sequencing, and Data Curation

This network enabled the linking of specific sequencing partners to specific sample acquisition sites, increasing sequencing volume in the state, shortening lines of communications, and providing surveillance coverage representative of all counties in North Carolina. Sample acquisition partners selected samples that were positive for SARS-CoV-2 on RT-PCR to send for sequencing. The primary goal was to select samples that were demographically (e.g., age, gender, race, ethnicity) and geographically representative of COVID-19 cases within the state.

Further, sample acquisition partners aimed to send samples from all COVID-19-positive hospitalizations and deaths, including pediatric deaths and cases of multisystem inflammatory syndrome in children (MIS-C). Partnering hospitals and sequencing centers directly minimized transport times and assured that there was no single point of failure, which was important early in the pandemic when supplies were erratic.

Adequate sampling coverage is crucial in detecting new variants when they emerge and accurately monitoring the prevalence of circulating variants (25). During the public health emergency phase of the pandemic, sequencing hubs aimed to maintain a sequencing volume of SARS-CoV-2-positive samples from sampling partners that amounted to ≥5% of reported statewide cases per week when COVID-19 case counts were >1,000 cases per week. During epidemic lulls where case counts were <100 cases per week, sequencing partners aimed to sequence 100% of samples they received that met sequencing requirements. Only samples with a cycle threshold (CT) ≤ 27, corresponding to high viral genome copy numbers, would be eligible for sequencing, as experience showed that low copy number samples yield incomplete genomes under standard, high throughput conditions (3). Samples meeting this requirement typically represent 10-40% of reported RT-PCR positive samples.

In addition to adequate sampling coverage and sampling volume, comprehensive near real-time surveillance depends on the timely turnaround of sequencing results. Sequencing partners regularly transmitted sequencing information to NC DHHS and uploaded de-identified versions to public databases, such as GISAID (26) and GenBank (27). Sequencing facilities aimed for a maximum turnaround time from positive sample identification to database upload of 14 days and a target turnaround time of ≤7 days. Only full-length, completed sequences that conformed with the GISAID or GenBank QC guidelines were submitted. Depending on the state of the pandemic, these represented between 20-80% of PCR-positive samples collected by sample acquisition partners. In addition to uploading sequence information to open-source sequence repositories, sequencing partners also submitted the following to NC DHHS: patient metadata (demographics, vaccination status, clinical outcomes), sampling date, sampling partner information, sequencing partner information, pangolin lineage, and GISAID and/or GenBank accession IDs.

Patient privacy and HIPAA compliance were key concerns due to linking PCR-positive test results to patient health data. Test results were reported to the NC DHHS, who acted as the honest broker to maintain patient privacy. Samples were de-identified before sequencing, and only non-identifying information was deposited publicly. The NC DHHS Division of Public Health meets the definition of a public health authority 42 C.F.R. § 164.501. Data collection for this project constituted a public health surveillance activity deemed not to be research under 45 CFR 46.102(l)(2).

Phylogenetic analysis was completed using Nextstrain (28) and epidemiologic analyses were completed in R (29). All CORVASEQ sequence data are available from GISAID (26) and Genbank (27) (accession numbers can be found in **Supplemental Data 1** within the manuscript’s Supporting Information files). Aggregate demographic information for CORVASEQ data can be found in **Supplemental Data 2**. Information on all reported SARS-CoV-2 cases in North Carolina (including demographics) is available from NCDHHS (https://covid19.ncdhhs.gov/dashboards/archived-data-behind-dashboards).

## Results

### Surveillance Metrics and Network Performance

The epidemiology data core was created to regularly assess multiple SARS-CoV-2 surveillance metrics reported by sequencing partners to NC DHHS. These metrics included SARS-CoV-2 variant profiles in the state, partner sequencing volume, turnaround time from sample acquisition to database upload, demographic and geographic trends in SARS-CoV-2 variant profiles, and reported vaccination status of individuals testing positive for COVID-19 whose samples were selected for sequencing. In addition to reporting regular metrics to NC DHHS, the epidemiology data core developed an interactive dashboard for the state to employ for SARS-CoV-2 surveillance and adapt to future pathogen surveillance needs. This dashboard provides information on SARS-CoV-2 and the CORVASEQ network aimed at the general public, including SARS-CoV-2 variant profiles over time (**Figure 2**) and by geographic region (**Figure 3**), demographic distribution of sequenced SARS-CoV-2 samples and all COVID-19 cases reported to NC DHHS (**Supplemental Figure 1**), and vaccination status and clinical outcomes (e.g., hospitalization, death) reported for individuals who had tested positive for SARS-CoV-2 and had their sample selected for sequencing.

**Figure 2.**
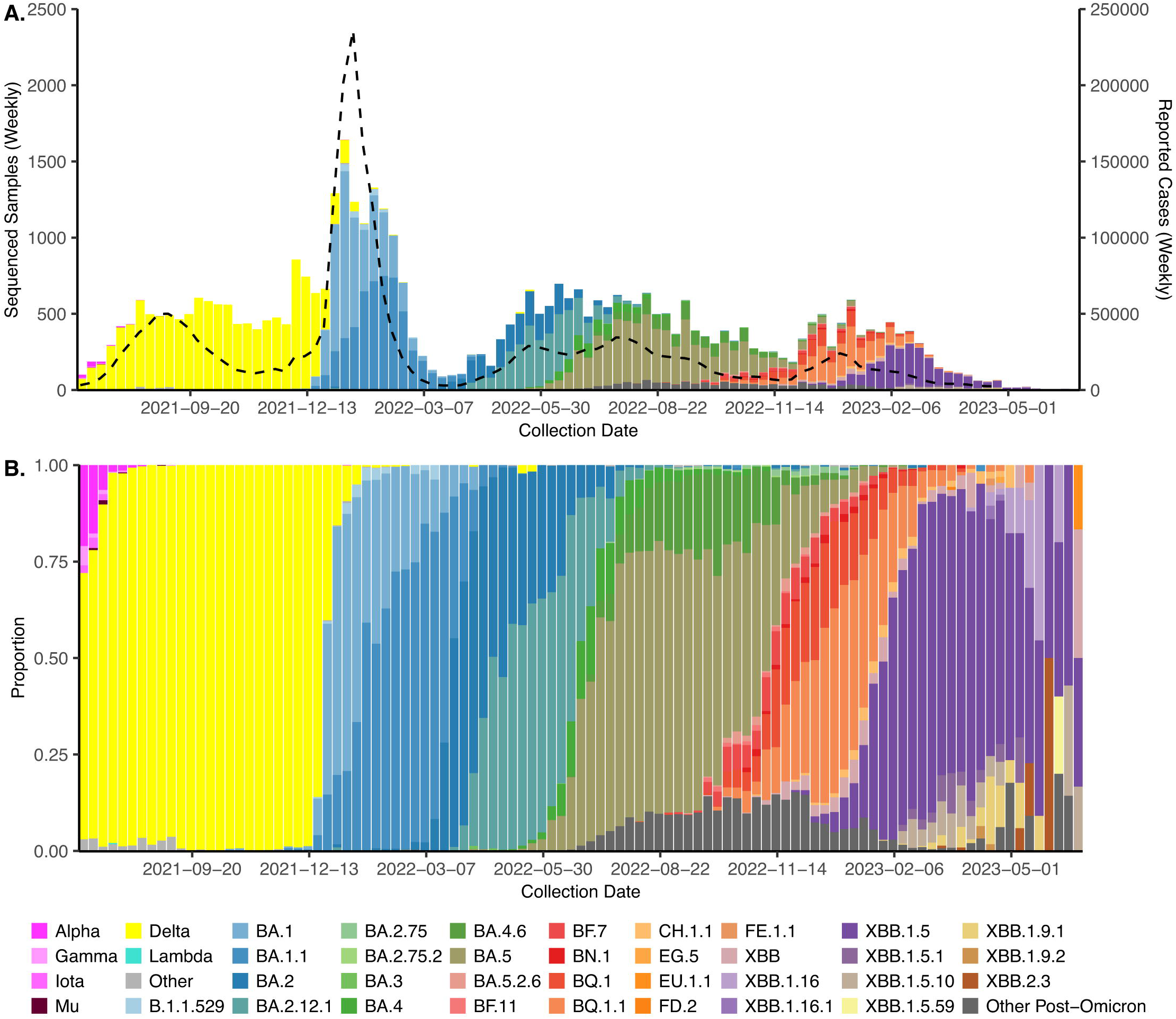
Sequenced Samples and Reported Cases. Weekly (**top**) sequencing volume shown (left axis) with reported cases (dashed line, right axis) and (**bottom**) variant profiles among sequences from the CORVASEQ network, May 2021 through June 2023, colored by variant.

**Figure 3.**
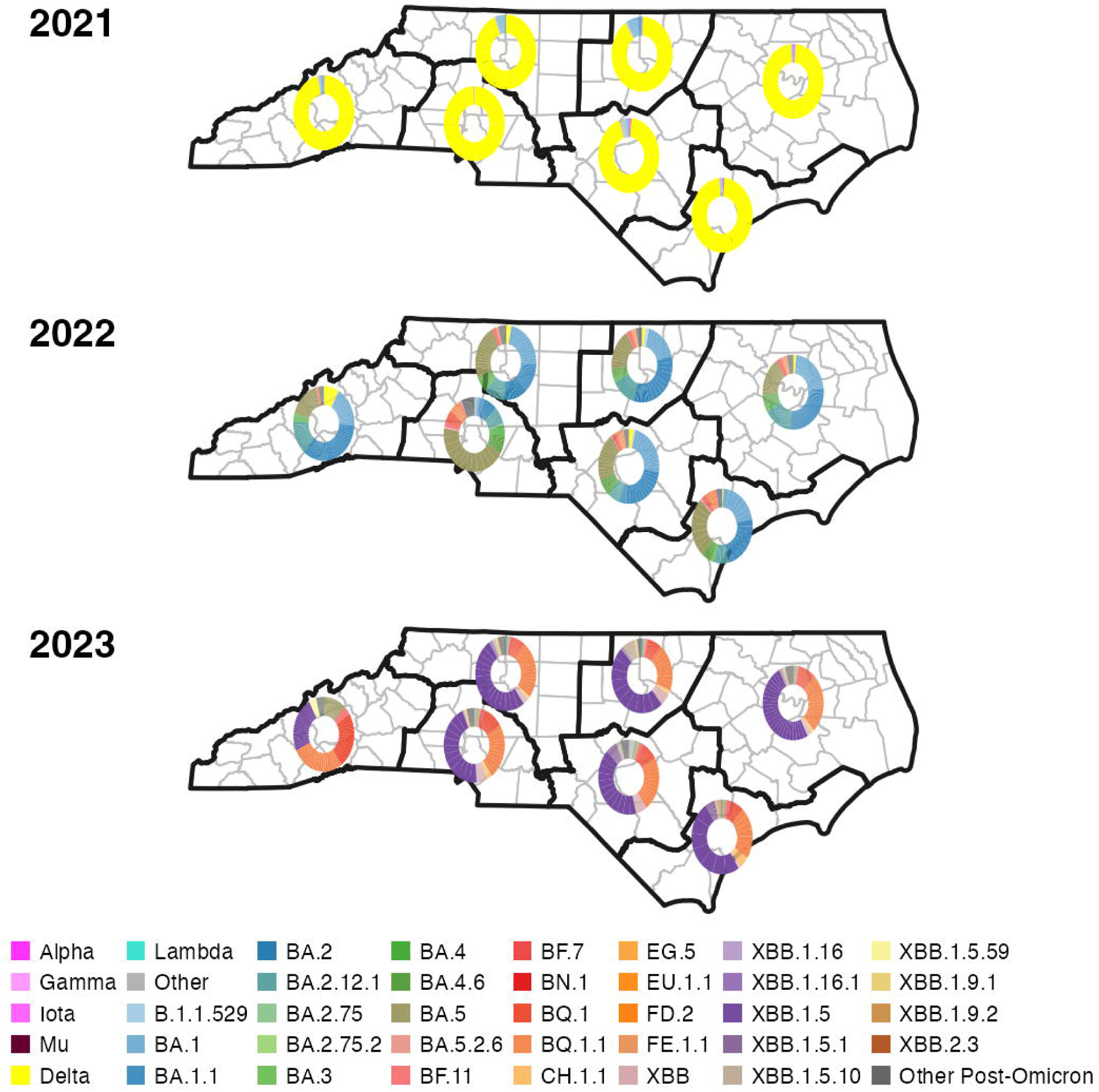
Geographic distribution of SARS-CoV-2 variants. Maps showing the geographic distribution of sequenced SARS-CoV-2 variants from CORVASEQ samples in North Carolina by flu region and year of sampling, 2021-2023.

CORVASEQ successfully met its goal of increasing viral sequencing volumes statewide. At peak volume during the height of the initial Omicron wave, CORVASEQ partners were sequencing over 500 samples per week statewide, with similar volumes achieved during the COVID-19 waves spurred by BA.2, BA.2.12.1, and BA.5 (**Figure 2**). By the time the federal COVID-19 public health emergency declaration expired in May 2023, the CORVASEQ network had contributed over 50,000 SARS-CoV-2 sequences from all 100 North Carolina counties to public databases. These samples selected for sequencing were demographically representative of reported COVID-19 cases (**S1 Figure**).

When the CORVASEQ network was first established, Delta was the dominant variant circulating in North Carolina and most of the world, consistently accounting for nearly 100% of weekly sequenced samples (**Figure 2**). When South African researchers discovered the highly transmissible Omicron variant in late November 2021 (4), the rest of the world was immediately on high alert to prepare for its spread. The CORVASEQ network detected the first known case of Omicron in North Carolina in early December 2021 (30), providing an early warning signal to NC DHHS about the arrival of this new divergent variant and allowing public health leaders to monitor viral dynamics and respond appropriately as Omicron quickly became the dominant SARS-CoV-2 variant in the state (**Figure 2**, **Figure 4**).

**Figure 4.**
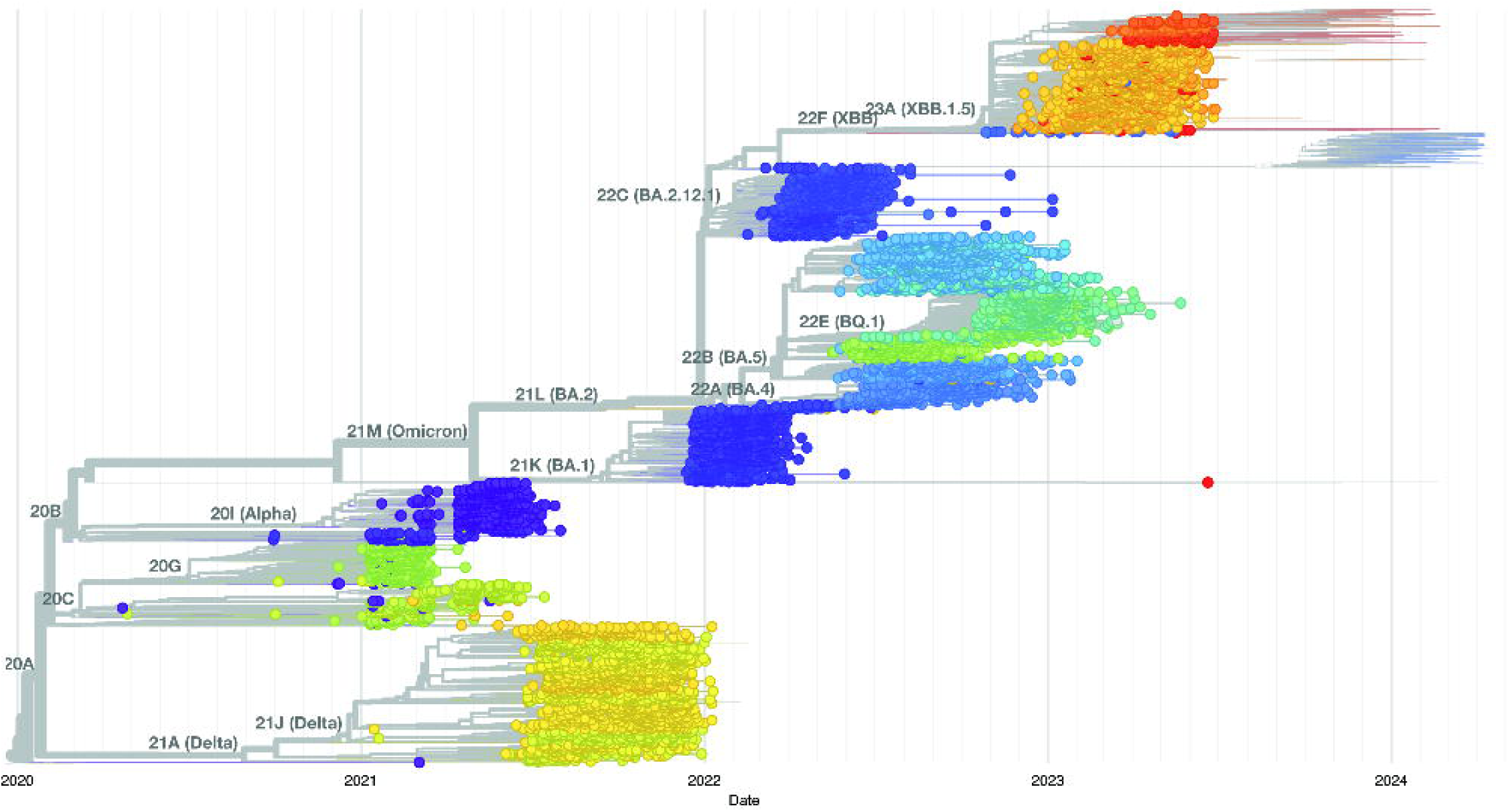
CORVASEQ-specific genomic surveillance build. Genomic build of SARS-CoV-2 sequences from CORVASEQ samples, 2021-2023.

Interestingly, with few exceptions, sequencing results across regions (**Figure 3**) in North Carolina showed similar distributions of SARS-CoV-2 variants each calendar year. This finding attests to the rapid spread of new variants across North Carolina, a geographically and demographically large and diverse state.

Throughout the public health emergency phase of the pandemic, this comprehensive network was a crucial resource for state public health officials, enabling communication about sequencing protocols and highlighting concerning signals in sequencing results. The arrival of Omicron evidenced the network’s capacity to quickly detect new variants (30). Continued sequencing efforts during the public health emergency proved vital in tracking the evolutionary dynamics of SARS-CoV-2 throughout the state and communicating the emergence of potentially concerning signals to the state, such as the detection of EG.5 in mid-April 2023 before it had even reached 0.5% prevalence nationally (**Figure 4**) (31).

## Discussion

### Readiness for Ongoing and Future Public Health Emergencies

The CORVASEQ network was borne of a need to establish timely and comprehensive genomic surveillance during an ongoing global health emergency. This need was recognized by state legislators, state and local public health officials, academic leadership, and CDC’s Epidemiology and Laboratory Capacity for Prevention and Control of Emerging Infectious Diseases (ELC) Program. Through the allocation of funds to build a comprehensive surveillance program, this partnership has brought together a multi-disciplinary team in the service of public health, including microbiologists, immunologists, epidemiologists, research computing specialists, project managers, medical professionals, and public health leaders to track an evolving pathogen in a large and diverse state. This coordinated network also sought to standardize and streamline pathogen genomic surveillance across institutions and establish standard modes of communication, allowing for rapid mobilization of resources in the event of future outbreaks.

By expanding genomic surveillance beyond tertiary hospitals to create a demographically and geographically representative surveillance system, new partnerships were formed between academic and non-academic healthcare and public health institutions that can continue to be leveraged to track SARS-CoV-2 and other pathogens. While no longer classified as a public health emergency, there is significant value in ongoing genomic surveillance of SARS-CoV-2, given the virus’ continued evolution and ensuing waves of COVID-19 outbreak. Despite the dramatic drop in sequences submitted to public databases since early 2023, genomic surveillance - in conjunction with other clinical and environmental surveillance metrics - continues to provide necessary information to public health professionals to plan for vaccine rollouts and tailor messaging around disease trends. Since the end of the formal public health emergency declaration, new sub-variants continue to emerge and change the landscape of SARS-CoV-2 infections in North Carolina and globally (31). As each new variant has risen to dominance, waves of increased infections, test positivity, wastewater signals, and hospitalizations have followed. The rich data from genomic surveillance networks like CORVASEQ allows for sophisticated analyses to understand pathogen transmission dynamics and evolution and retrospective analyses of sampling strategies to improve surveillance system design (25).

The utility of the CORVASEQ network and tools developed through this partnership is not limited to genomic surveillance of SARS-CoV-2; it can also be leveraged for surveillance of seasonal outbreaks, such as RSV and seasonal influenza in a similar manner to VISION and other national networks, and in future public health emergencies. As it is price-prohibitive for most jurisdictions to maintain high-level, high-detail, rapid-turnaround capacity all the time, partnership-based networks that can be operationalized quickly provide one alternative for future pandemic outbreaks (32). In the few years since SARS-CoV-2 emerged, there have been threats from multiple newly emerging or re-emerging pathogens that could require access to rapid, representative genomic sequencing data. Mpox, first identified as a cause of sporadic outbreaks in humans in 1970, began increasing in incidence in recent decades while remaining geographically confined to the African continent (33). In 2022, mpox began showing sustained human-to-human transmission, particularly among men who have sex with men (MSM), followed by rapid global spread, with phylogenetic analyses showing clear evidence of viral evolution and distinct circulating strains. Genomic surveillance networks can also be deployed to monitor recent increases in vector-borne infections globally and in the US, such as local outbreaks of dengue virus in Florida in recent years (34). Finally, highly pathogenic avian influenza A (HPAI A) H5N1 is an ongoing public health threat that will require a coordinated and quick response that is adaptable to the rapidly evolving disease landscape. HPAI A H5N1 first spread to wild birds in North America in 2021 (35), followed by large outbreaks in wild birds, commercial poultry, and multiple species of mammals (36,37). Since March 2024, large outbreaks in dairy cattle have been discovered in several US states, along with the first documented case of cow-to-human transmission (38–40). As the situation with HPAI H5N1 continues to unfold, mobilization of existing surveillance networks such as CORVASEQ and VISION, as well as veterinary surveillance networks and genomic data from wastewater surveillance, will be critical in monitoring disease spread in animals and timely detection of human cases that may emerge.

### Lessons Learned

While the success of CORVASEQ is a testament to the strength of North Carolina’s academic, clinical, and public health institutions, these qualities are not unique to the CORVASEQ member institutions. The CORVASEQ design should be considered as a model for other jurisdictions to maximize surveillance capabilities. A major achievement resulting from the creation of this network was the facilitation of regular communication between NC DHHS, hospital systems, and academic institutions. In order to be maximally effective, public health cannot be siloed; these types of partnerships and strong connectivity between the parties involved are essential.

There were challenges faced during the conceptualization and creation of this network that are important to understand and improve upon in future work. A key, but particularly time-consuming, logistical step in creating the network was the drafting and execution of contracts and data use agreements between CORVASEQ partners and state agencies. Drafting budgets and ensuring funds were dispersed in a timely fashion to all partners was critical in ensuring that sequencing efforts could be ramped up in a timely manner. Additionally, developing and implementing common formatting, naming conventions, and data transmission processes required clear documentation, communication, and buy-in from all parties. The common theme in the challenges faced was the amount of time and resources required to get the network up and running; because the logistics involved in creating a network can be time-consuming, the optimal time to build the foundation for these types of networks is during times of relative calm (as opposed to during an outbreak). These logistics include identifying partners, drafting contracts and data use agreements, developing laboratory surveillance protocols, and opening lines of communication. Given the challenges faced with establishing electronic laboratory reporting (ELR) at CORVASEQ partner sites, there is a need for improved ease of transfer of PHI, lab results, and metadata to health departments. These preparedness steps are critical hurdles to overcome early so that in times of crises, networks can be rapidly mobilized. Once established, a key consideration is maintaining the necessary infrastructure and open lines of communication so that the network can be quickly revived in an emergency. Specifically, this includes perpetual data use agreements for data sharing in public health emergencies, draft contracts that can be amended rapidly, malleable software that can easily accommodate the transmission of new data, and ongoing relationships between stakeholders in local health systems and health departments who can quickly scale systems as needed to accommodate the needs introduced by a new public health threat. An example of this in practice is the newly established Insight Net, established in 2023 by the CDC’s Center for Forecasting and Outbreak Analytics. Insight Net is a network of 13 partner centers representing over 100 academic and private institutions that focus on improving the capacity to anticipate and respond to disease outbreaks before a public health emergency occurs.

## Conclusions

In conclusion, the establishment of CORVASEQ was successful in developing a collaborative and comprehensive pathogen genomic sequencing network to monitor SARS-CoV-2 as the virus evolved and spread in North Carolina during the public health emergency phase of the pandemic. This effort resulted in standardized statewide protocols for pathogen genomic surveillance, a comprehensive dashboard that can be adapted to various pathogens, and open channels of communication between sample acquisition partners, sequencing facilities and academic institutions, and public health partners at the county and state level. The models and tools developed for the CORVASEQ network will allow us to rapidly mobilize in response to ongoing and emerging public health threats in North Carolina. The successes from CORVASEQ, taken together with lessons learned along the way, can serve as a useful roadmap for other states or areas as they seek to build or enhance pathogen genomic surveillance networks.

## Data Availability

All CORVASEQ sequence data are available from GISAID (26) and Genbank (27) (accession numbers can be found in S1 Data within the manuscript's Supporting Information files). Aggregate demographic information for CORVASEQ data can be found in S2 Data. Information on all reported SARS-CoV-2 cases in North Carolina (including demographics) is available from NCDHHS (https://covid19.ncdhhs.gov/dashboards/archived-data-behind-dashboards).

## Acknowledgments

We acknowledge additional support from the UNC Lineberger Comprehensive Cancer Center and the UNC School of Medicine. None of this would have been possible without our dedicated staff doing the infectious sample collection, processing, sequencing, and analysis: Linda J. Pluta, Patricio Cano, Anjelica Juarez, Razia Moorad, Justin Landis, Paul Oglesby Grant Greenwood, Caitlin Edwards, Sinead Isaacson, and Sayal Guirales-Medrano. At ECU Health and Brody School of Medicine, we acknowledge Heather Duncan and Kim Briley for specimen collection from all counties in Eastern North Carolina (ENC) and Weihua Huang and Changhong Yin for processing and sequencing ENC samples as well as GISAID submissions. We would like to acknowledge Catherine Passaretti MD, Michael DeWitt, Jessica Kearney-Bryan, Amanda Heiliger, and the Atrium Health Internal Medicine research and laboratory teams for supporting the sequencing efforts at Atrium Health. At Wake Forest University School of Medicine, we would like to thank Kara Libby, Tara Staley, Michaela Romero, Tracey Young, Douglas McGlasson, and Michael DeWitt for their efforts in support of this project. At Duke University, we want to acknowledge the following dedicated faculty, staff, and clinicians: Christopher Polage, Kari Ryan, Steve Haase, Tony Moody, Thad Gurley, Cassie Porth, Darin Weed, Alejandro Berrio Escobar, and Devjanee Swain-Lenz, who synergistically worked together to do the sample collections, processing, logistical coordination, sequencing, and data analysis.

Additionally, we would like to thank Chris Woods and Brad Nicholson for the collection and coordination of samples from Veterans Affairs Medical Centers located in North Carolina to Duke Center for Genomic Computation Biology (GCG) for sequencing.

## Supporting Information

**Supplemental Figure 1. Demographic distribution of CORVASEQ samples selected for sequencing and COVID-19 cases reported to NCDHHS.** Weekly demographic distribution of cases reported to NCDHHS (top) and CORVASEQ samples (bottom) by (**A**) sex, (**B**) age, (**C**) race, and (**D**) ethnicity.

**Supplemental Data 1. Accession numbers for CORVASEQ samples.**

**Supplemental Data 2. Aggregate counts of CORVASEQ samples by demographic characteristics.**

