## Supplementary figures and images for "North Carolina CORonavirus VAriant SEQuencing (CORVASEQ): A surveillance network to monitor an evolving pandemic"

### Supplemental Figure 1

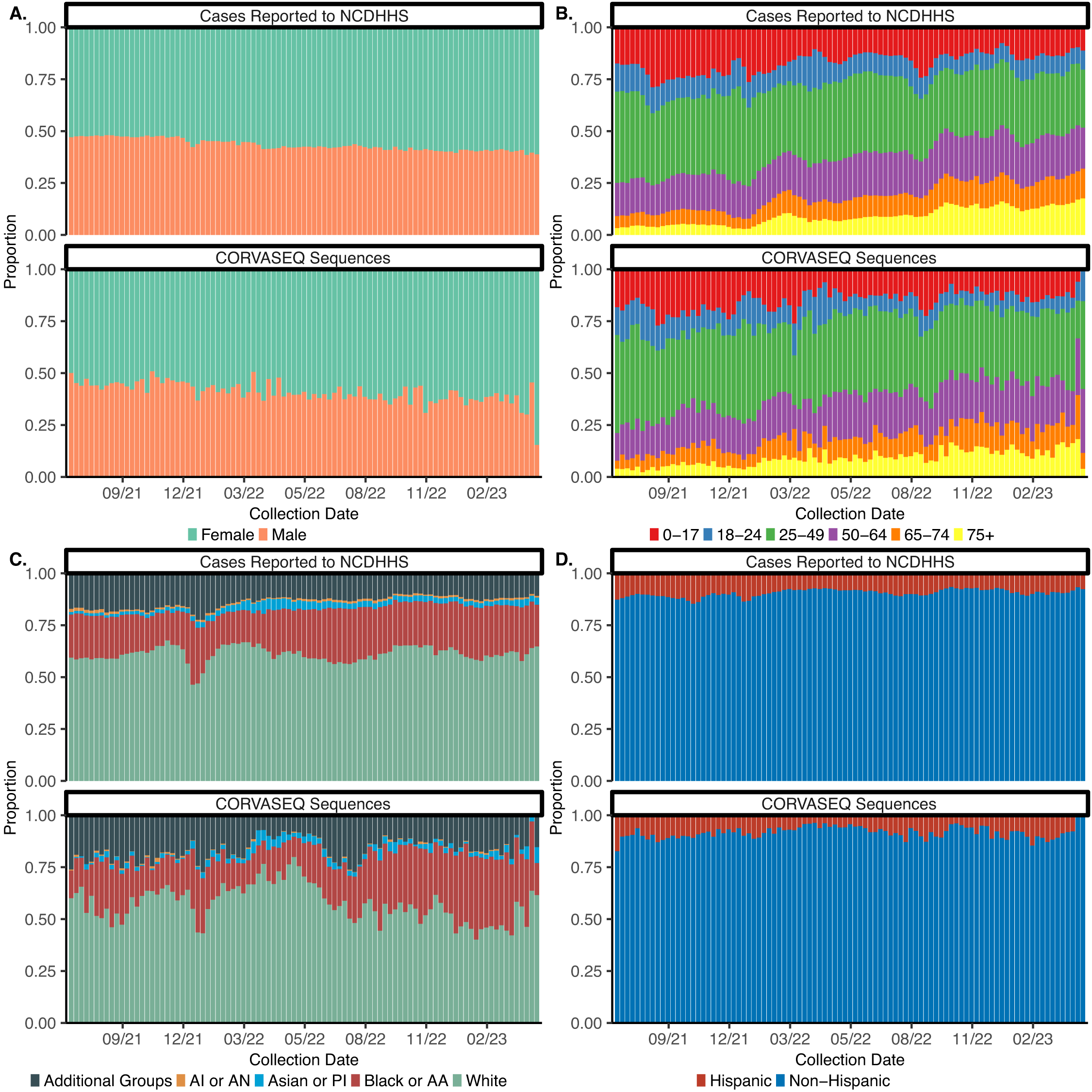
